# Heterogeneity of the Effect of Telemedicine Hypertension Management Approach on Blood Pressure: A Systematic Review and Meta-analysis of US-based Clinical Trials

**DOI:** 10.1101/2023.09.14.23295587

**Authors:** Sameer Acharya, Gagan Neupane, Austin Seals, KC Madhav, Dean Giustini, Sharan Sharma, Yhenneko J. Taylor, Deepak Palakshappa, Jeff D. Williamson, Justin B. Moore, Hayden B. Bosworth, Yashashwi Pokharel

## Abstract

**Background:** Telemedicine management of hypertension (TM-HTN) uses home blood pressure (BP) to guide pharmacotherapy and telemedicine-based self-management support (SMS). Optimal approach to implementing TM-HTN in the US is unknown.

**Methods:** We conducted a systematic review and a meta-analysis to examine the effect of TM-HTN vs. usual clinic-based care on BP and assessed heterogeneity by patient- and clinician-related factors. We searched US-based randomized clinical trials among adults from Medline, Embase, CENTRAL, CINAHL, PsycInfo, and Compendex, Web of Science Core Collection, Scopus, and two trial registries to 7/7/2023. Two authors extracted, and a third author confirmed data. We used trial-level differences in systolic BP (SBP), diastolic BP (DBP) and BP control rate at ≥6 months using random-effects models. We examined heterogeneity of effect in univariable meta-regression and in pre-specified subgroups [clinicians leading pharmacotherapy (physician vs. non-physician), SMS (pharmacist vs. nurse), White vs. non-White patient predominant trials (>50% patients/trial), diabetes predominant trials (≥25% patients/trial) and in trials that have majority of both non-White patients and patients with diabetes vs. White patient predominant but not diabetes predominant trials.

**Results:** Thirteen, 11 and 7 trials were eligible for SBP, DBP and BP control, respectively. Differences in SBP, DBP and BP control rate were -7.3 mmHg (95% CI: - 9.4, -5.2), -2.7 mmHg (-4.0, -1.5) and 10.1% (0.4%, 19.9%), respectively, favoring TM-HTN. More BP reduction occurred in trials with non-physician vs. physician led pharmacotherapy (9.3/4.0 mmHg vs. 4.9/1.1 mmHg, P<0.01 for both SBP/DBP), pharmacist vs. nurses provided SMS (9.3/4.1 mmHg vs. 5.6/1.0 mmHg, P=0.01 for SBP, P<0.01 for DBP), and White vs. non-White patient predominant trials (9.3/4.0 mmHg vs. 4.4/1.1 mmHg, P<0.01 for both SBP/DBP), with no difference by diabetes predominant trials. Lower BP reduction occurred in both diabetes and non-White patient predominant trials vs. White patient predominant but not diabetes predominant trials (4.5/0.9 mmHg vs. 9.5/4.2 mmHg, P<0.01 for both SBP/DBP).

**Conclusions:** TM-HTN is more effective than clinic-based care in the US, particularly when non-physician led pharmacotherapy and pharmacist provided SMS. Non-White patient predominant trials seemed to achieve lesser BP reduction. Equity conscious, locally informed adaptation of TM-HTN is needed before wider implementation.

**Clinical Perspective:** *What Is New?:*

- In this systematic review and meta-analysis of US-based clinical trials, we found that telemedicine management of hypertension (TM-HTN) was more effective in reducing and controlling blood pressure (BP) compared with clinic based hypertension (HTN) care.
- The BP reduction was more evident when pharmacotherapy was led by non-physician compared with physicians and HTN self-management support was provided by clinical pharmacists compared with nurses,
- Non-White patient predominant trials achieved lesser BP reductions than White patient predominant trials.

*What Are the Clinical Implications?:*

- Before wider implementation of TM-HTN intervention in the US, locally informed adaptation, such as optimizing the team-based HTN care approach, can provide more effective BP control.
- Without equity focused tailoring, TM-HTN intervention implemented as such can exacerbate inequities in BP control among non-White patients in the US.

## Introduction

Approximately 47% of American adults have hypertension (HTN) but only a quarter of them achieve blood pressure (BP) control, with worse control among non-Hispanic Black or Hispanic adults than White Americans.^1^ Managing HTN using traditional clinic-based model has several challenges.^2^ Assessment of BP during brief visits are often inaccurate.^3^ To confidently assess accurate BP control in clinic, it may require up to 5-6 BP measurements,^4^ which is not realistic in routine practice. Self-measurement of BP by patients offers more BP data and improves clinicians’ confidence about accuracy in BP control.^5,6^ Furthermore, enhancing patients’ HTN self-care skills is important,^7^ which is often difficult within the scope of a typical clinic visit. Telemedicine management of hypertension (TM-HTN) employs a focused team-based approach, where pharmacotherapy is based on self-measured BP (SMBP). Ideally, SMBP is transmitted using a telemonitoring platform, and self-management support (SMS) is offered via telemedicine.^2^ While TM-HTN has the promise to overcome the challenges of clinic-based HTN care, understanding how patient and clinician-related factors influence TM-HTN’s effect on BP is important.

Prior meta-analyses including data outside the US have shown that SMBP with additional support can improve BP.^8-10^ The additional support included heterogeneous levels of SMS with or without SMBP-guided pharmacotherapy. SMBP-guided pharmacotherapy is the essential core of TM-HTN. To our knowledge, we are not aware of any meta-analysis limited to SMBP-guided pharmacotherapy. Accordingly, we conducted a systematic review and meta-analysis of US-based trials that used SMBP-guided pharmacotherapy. Local US-based data is particularly relevant to inform TM-HTN implementation because of the global differences in health care systems and practices. Little is known whether patient and clinician-related factors in the US influence TM-HTN’s effect on BP. Generally, BP control is better when medication prescription for HTN is led by non-physicians than by physicians,^11,12^ but it is unknown whether such heterogeneity exists with TM-HTN. Previous meta-analyses have examined the effect of various implementation strategies on BP, with team-based approach having the highest impact. This approach involves task shifting or sharing responsibilities to non-physicians like pharmacists and nurses.^11-13^ As a team-based approach in TM-HTN, either physicians or non-physicians could deliver SMBP-guided pharmacotherapy.^2^ It is unknown whether BP achieved in a TM-HTN program varies when the pharmacotherapy is led by physicians vs. non-physicians, or when SMS is provided by different clinicians. Similarly, whether TM-HTN’s effect varies by patient characteristics is unknown. For example, although Black and non-Black patients derive similar benefits from BP control,^14^ there is a persistent disparity in BP control in Black compared with White patients.^1^ Therefore, implementing TM-HTN without purposeful equity-conscious delivery plan has the risk of exacerbating inequities in BP control. Accordingly, using a systematic review and meta-analysis of US-based clinical trials of TM-HTN, we examined the effect of TM-HTN vs. usual care on difference in systolic BP (SBP), diastolic BP (DBP) and BP control rate and the heterogeneity of TM-HTN’s effect by various patient and clinician characteristics.

## Methods

We registered our study protocol on PROSPERO (CRD42022324076) and Open Science Framework.^15^

### Data Sources and Searches

Together with an academic health sciences librarian, we developed and used a search strategy in six bibliographic databases till July 7, 2023 [Medline (Ovid), Embase (Ovid), CENTRAL (Ovid), CINAHL (Ebsco), PsycInfo (Ebsco), and Compendex]; two citation indexes (Web of Science Core Collection and Scopus) and trial registries (ClinicalTrials.Gov and International Clinical Trials Registry Platform). Several definitions of telemedicine informed our searches.^2,16,17^ We examined broader telemedicine and telehealth terminologies, without limits or exclusions, used in the remote monitoring of HTN and BP (Supplement Table 1). We created a list of search terms using exploratory searches and key seed papers, to fit the review’s scope:^18^ telemedicine and telehealth strategies (e.g., mobile health, telehealth, telemonitoring, telemedicine, mHealth, eHealth, videoconferencing, smartphones, virtual care, remote monitoring, health information technology) combined with home BP monitoring (SMBP, home BP measurement, ambulatory BP) and United States-related geographic filter.^19^ We limited our searches to randomized controlled trials. We obtained a peer review of the Medline search strategy from an information specialist from Cochrane HTN.

### Trial Selection

The essential trial selection required that the clinical trial be US-based, intervention used SMBP-guided pharmacotherapy, and SMS was optional. Trials that used SMBP without SMBP-guided pharmacotherapy were excluded. Usual care group used clinic-based HTN management without a telemedicine program where pharmacotherapy was not guided by SMBP. Other eligibility criteria are available in the study protocol.

### Data Extraction and Quality Assessment

We saved search results in EndNote, removed duplicate references and transferred the citations into Covidence. Two reviewers (S.A and G.N) independently performed title, abstract, and full-text screening and data abstraction. A third reviewer (Y.P) resolved any disagreements and confirmed trials eligibility and data.

When there were more than one comparison arms present in a trial, we used the data from the arm with the most intense intervention as appropriate to our research question. For example, with three arms, 1) usual care, 2) SMBP, and 3) SMBP plus SMS we presented the results for the comparison between 1) usual care, and 3) SMBP plus SMS.

We collected trial level data on patient’s mean age, sex, race, education, baseline SBP and DBP, clinicians leading pharmacotherapy and SMS, trial publication year and TM-HTN intervention duration. We found that the clinicians leading pharmacotherapy were either physicians, pharmacists or nurses. Similarly, the clinicians providing SMS were either pharmacists or nurses. We further defined clinicians leading pharmacotherapy (physicians vs. non-physicians) and SMS (pharmacists vs. nurses). If a non-physician (e.g., pharmacist) prescribed anti-hypertensive medications without routine and direct approval by physician, then we considered the pharmacotherapy to be led by the pharmacist.^12^ Conversely, if the pharmacist prescribed the medications but required routine and direct approval by physician, we considered the pharmacotherapy to be led by physician. We defined White vs. non-White patient predominant trials (>50% White or non-White patients per trial) and diabetes predominant trials (≥25% vs. <25% of participants with diabetes per trial).

Two authors (M.K. and Y.P.) independently assessed the quality of included trials using the Cochrane risk of bias (RoB) tool version 6.3 and resolved any differences by consensus.^20^ Two authors (M.K. and Y.P.) independently assessed the strength of evidence using the AHRQ criteria and resolved any differences by consensus.^21^ Supplement Table 2 shows PRISMA checklist.

### Data Synthesis and Statistical Analysis

Our outcomes of interest were difference in SBP, DBP and trial defined BP control rate between TM-HTN and usual clinic-based care at ≥6 months of the intervention.^14^ If multiple BP data were reported within a trial, we used the data for which the sample size was powered for. For example, if the trial provided BP data at 6, 12 and 18 months, but the trial was powered for 12-month BP data, we considered 12-month data for our analysis. We used random effects models, with Hartung-Knapp adjustment for between study heterogeneity, to examine the difference in BP outcomes between intervention and usual care. Throughout the study design, we ensured randomized comparisons between TM-HTN and usual care as conducted in individual trials. We calculated standard errors for each trial’s effect size from the reported confidence intervals or P-value. We conducted analyses in R (4.0.2) using the packages “meta”, “metafor” and “dmetar”. We considered two-sided P value <0.05 to be statistically significant and did not adjust for multiple comparisons. We quantified between-study heterogeneity using *I*^2^ statistic. We used a comparison-adjusted funnel plot with Egger’s test to assess for publication bias. We performed leave one out analysis to estimate the influence of each trial on the pooled effect.

We first examined the association of several patient and clinician-related variables with SBP and DBP difference in univariable meta-regression. These variables include patients’ mean age, sex (percentage of women/trial), race (White vs. non-White patient predominant trials), education (percentage of participants/trial with ≤high school education), baseline SBP or DBP, diabetes (diabetes patient predominant trials), clinicians leading pharmacotherapy (physicians vs. non-physicians) and SMS (pharmacists vs. nurses) and TM-HTN intervention duration. If there was a significant association for any of these variables, we considered sub-group analysis. As defined priori, we also performed sub-group analyses based on the clinician leading pharmacotherapy and SMS, White vs. non-White patient predominant trials, diabetes predominant vs. non-dominant trials and in trials that have majority of both non-White patients and patients with diabetes vs. White patient predominant but not diabetes predominant trials.

We identified trials with well-developed SMS focused on enhancing patient’s self-care skills. Such trials required that SMS was tailored for individual patients plus had a considerable emphasis on SMS assessed by frequency, content, and duration of contact.^22^ We further noted home BP data was transferred to clinics or investigators either by mail or telemonitoring device. To provide insights into using contemporary TM-HTN programs, in sensitivity analysis, we repeated analysis limiting to trials that used telemonitoring for SMBP.

## Results

### Trial Characteristics

We identified 13 eligible trials for SBP, 11 trials for DBP, and 7 trials for BP control, published between 1992-2019 in the US, with 5,330, 4,243 and 3766 participants, respectively (Figure 1).^23-35^ Two trials had cluster randomized design^23,28^ and others randomized individual participants (Table 1). TM-HTN intervention was leveraged via community in two trials, where patients’ physicians used SMBP to guide pharmacotherapy.^23,30^ Usual care in all trials comprised clinic-based HTN management with options for 1-2 pharmacist visits in one trial,^28^ and text-messaging encouraging lifestyle change in another trial.^34^ Supplement Tables 3-4 provide additional information about trials’ eligibility, intervention, procedures, and outcomes. Across all trials, mean (SD) age was 60.2 (5.4) years, 47.5% (21.9) were women, 38.2% (23.5) had ≤high school education, baseline SBP and DBP were 147.4 (8.6) mmHg and 86.6 (3.5) mmHg, respectively. The proportion of non-White patients ranged from 4-100% per trial. Seven were non-White patient predominant trials (four had >50% and other two trials had 39-48% Black participants/trial; one trial was exclusively in Hispanic adults). Similarly, the proportion of patients with diabetes per trial ranged from 14-100%, and eight were diabetes patient predominant trials (five were both non-White and diabetes patient predominant trials; three were both White and diabetes patient predominant trials). None of the non-White patient predominant trials had pharmacotherapy led by non-physicians or SMS provided by pharmacists. Seven trials had well-developed SMS and nine trials used telemonitoring of SMBP (four trials asked patients to mail home BP log) (Table 1 and Supplement Table 4). Well-developed SMS was present in two White and five non-White patient predominant trials, respectively.

**Figure 1.**
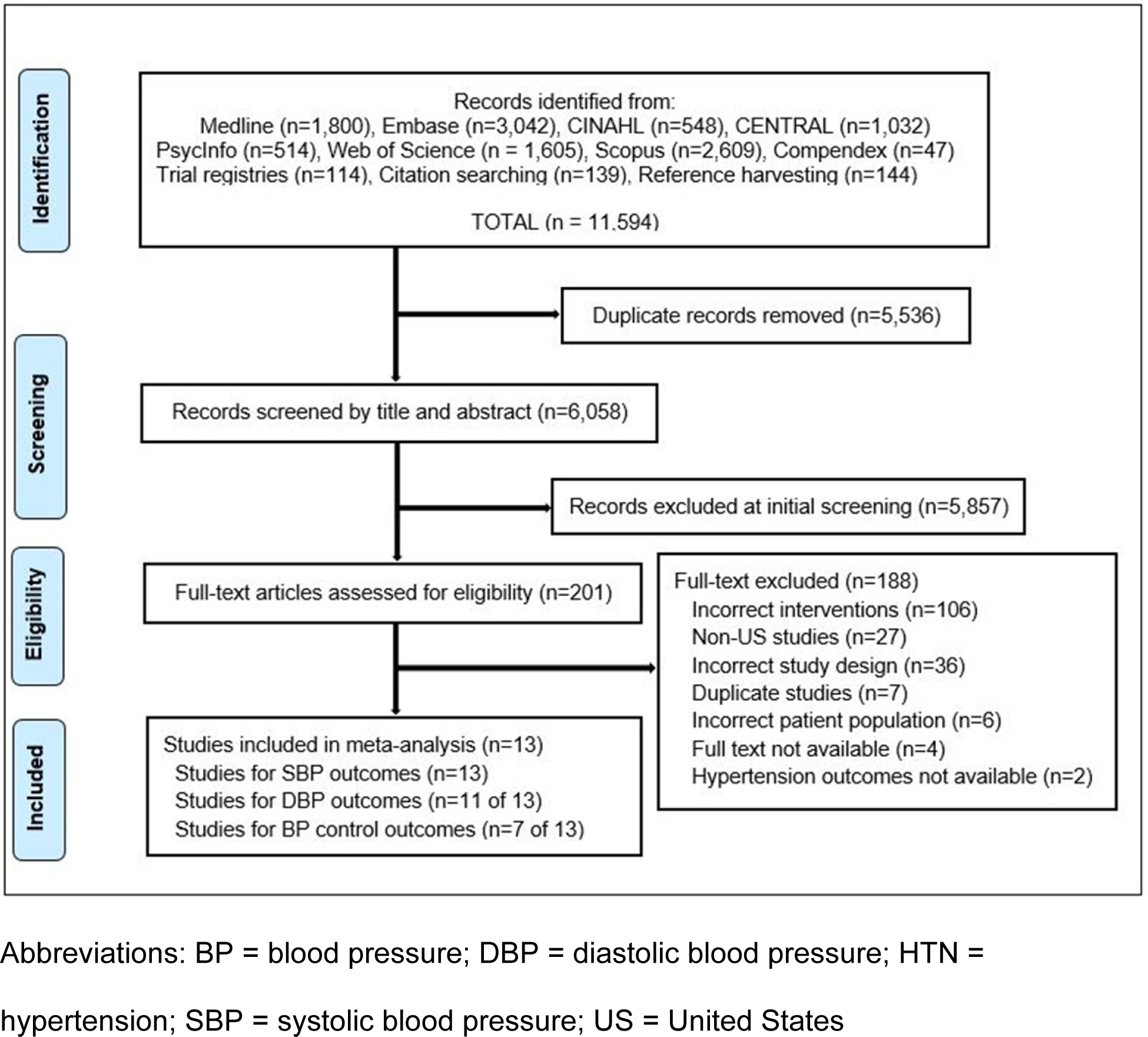
PRISMA flow diagram of search results and eligible articles.

**Table 1.**
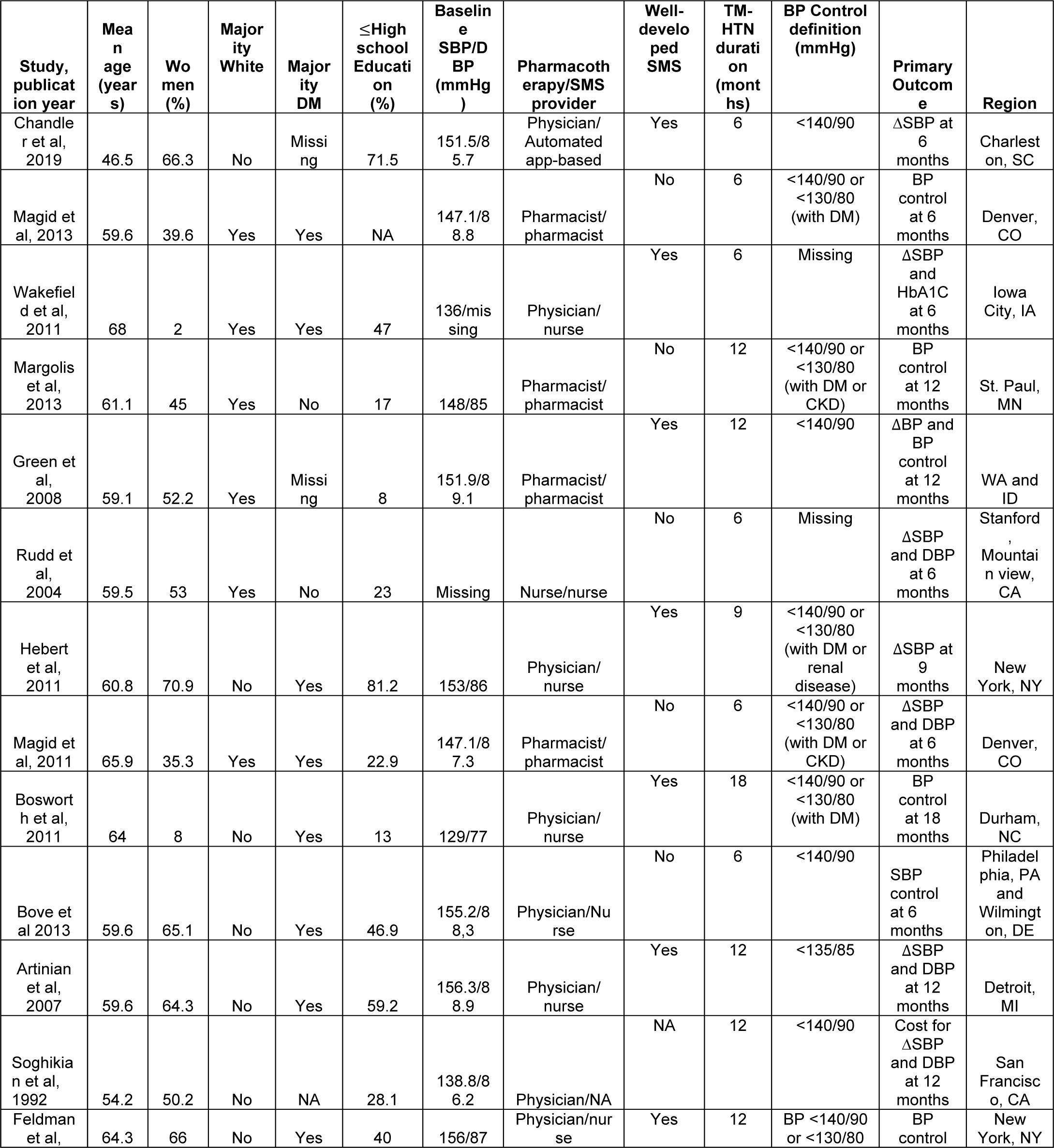
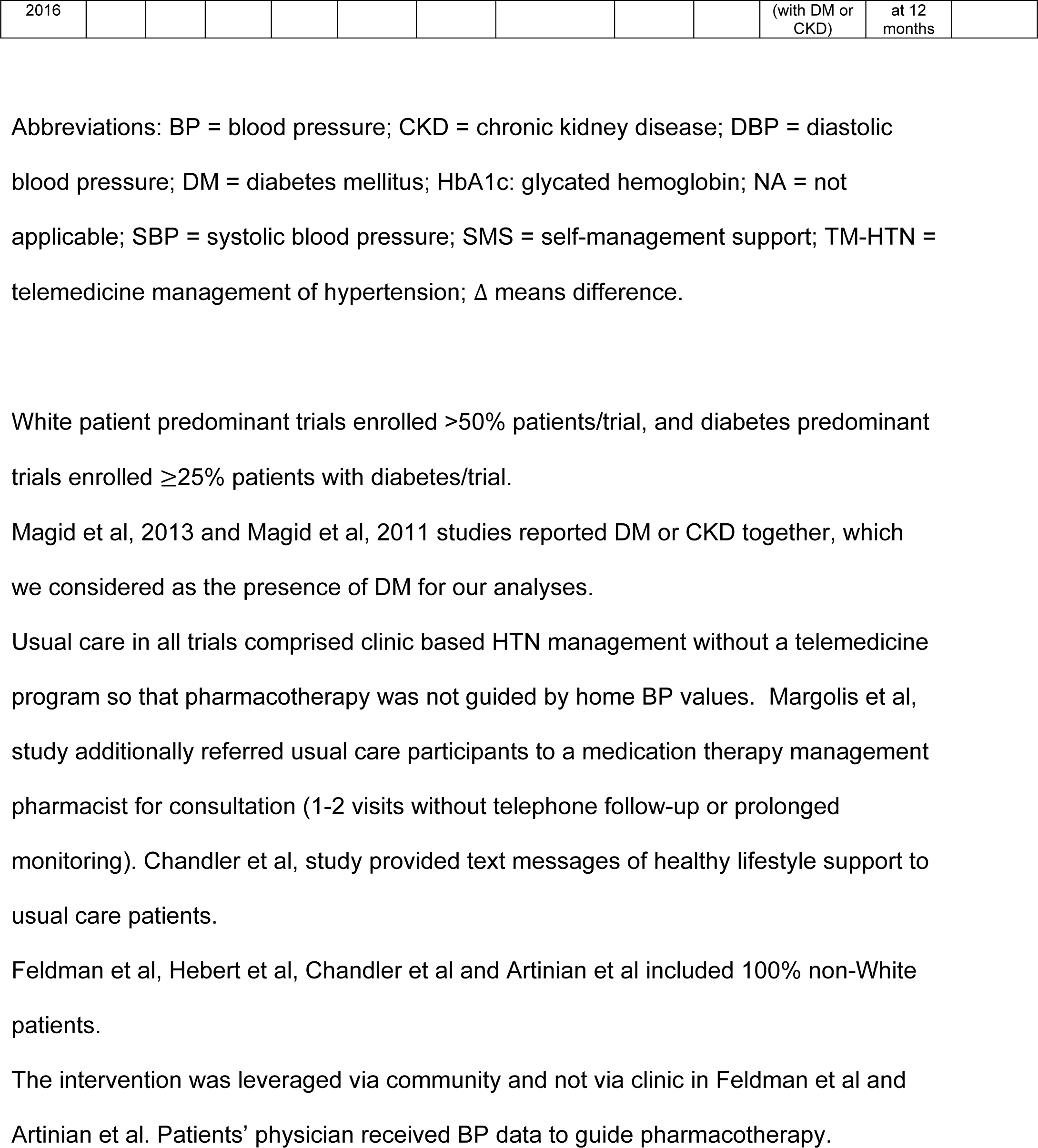
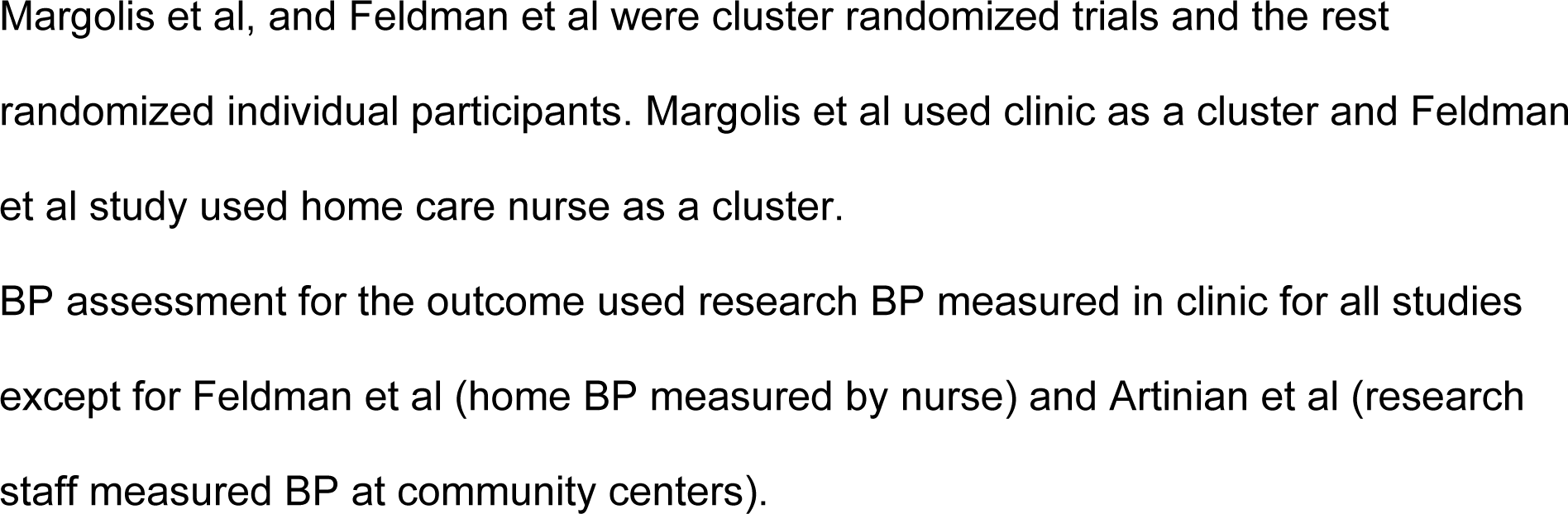
Key features of included trials.

Three trials (published between 1992-2007) had “some concerns” in RoB assessment from lack of pre-specified protocols.^29,30,33^ One trial had high RoB from large missing outcome data (31.2%).^23^ Others had low RoB (Supplement Figure 1). We did not find publication bias (Egger test P=0.83 and 0.26 for SBP and DBP, respectively, Supplement Figure 2). The *I*^2^ statistic was 53% (P=0.01) and 70% (P<0.01) for SBP and DBP, respectively, suggesting some heterogeneity among trials.^36^ Magid et al, 2013, Soghikian et al, and Artinian et al contributed the most to the heterogeneity in BP difference (Supplement Figure 3). We found high strength of evidence for both SBP and DBP difference (Supplement Table 5).

### BP Outcomes

The pooled SBP difference was -7.3 mmHg (95% CI: -9.4, -5.2), favoring TM-HTN intervention (Figure 2). In univariable meta-regression, the only variables significantly associated with SBP difference were clinicians leading pharmacotherapy and race. SBP reduction was 4.4 mmHg more when pharmacotherapy was led by non-physicians vs. physicians (P<0.01). For every 10% more non-White patients per trial, SBP difference achieved was 0.6 mmHg lower (P=0.03) or 5.0 mmHg lower SBP reduction in non-White vs. White patient predominant trials (P<0.01). In subgroup analyses, SBP reductions were -9.3 mmHg (95% CI: -11.9, -6.6) vs. -4.9 mmHg (95% CI: -7.3, -2.5) when pharmacotherapy was led by non-physician vs. physician (P<0.01), and -5.6 mmHg (95% CI: -7.8, -3.3) vs. -9.3 mmHg (95% CI: -13.1, -5.6) when SMS was provided by nurses vs. pharmacists (P=0.01), respectively, all favoring TM-HTN (Figures 3-4). Similarly, SBP reductions were -4.4 mmHg (95% CI: -6.6, -2.2) vs. -9.3 mmHg (95% CI: -11.4, -7.2) in non-White vs. White patient predominant trials (P<0.01), respectively, favoring TM-HTN (Figure 5). There were no significant subgroup differences by diabetes predominant trials (P=0.14). Both diabetes and non-White patient predominant trials achieved -4.5 mmHg (95% CI: -5.9, -3.1) SBP reduction vs. - 9.6 mmHg (95% CI: -12.7, -6.4) in White patient predominant but not diabetes predominant trials (P<0.01), favoring TM-HTN.

**Figure 2:**
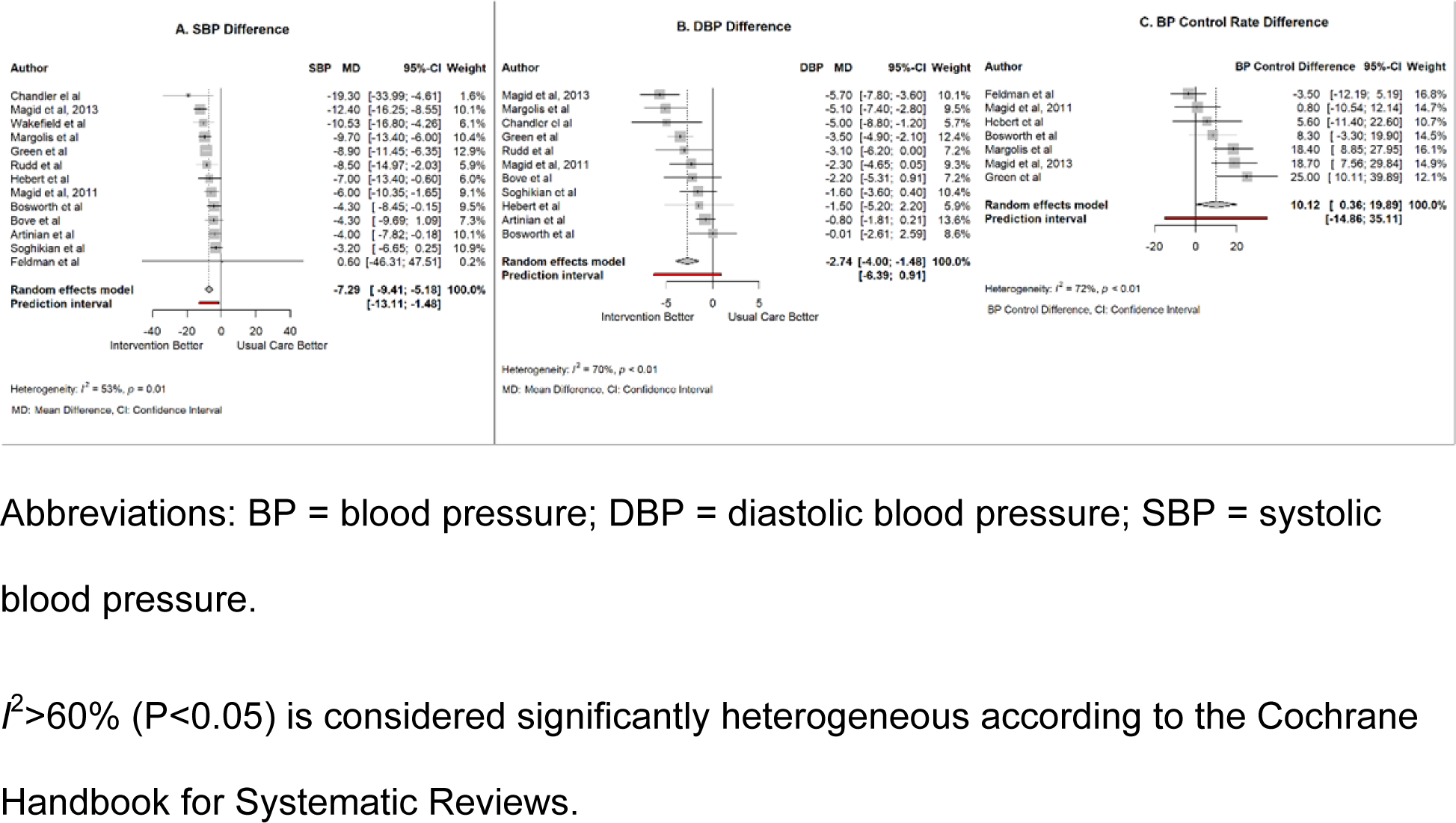
Mean overall pooled difference in SBP, DBP and BP control rate.

**Figure 3:**
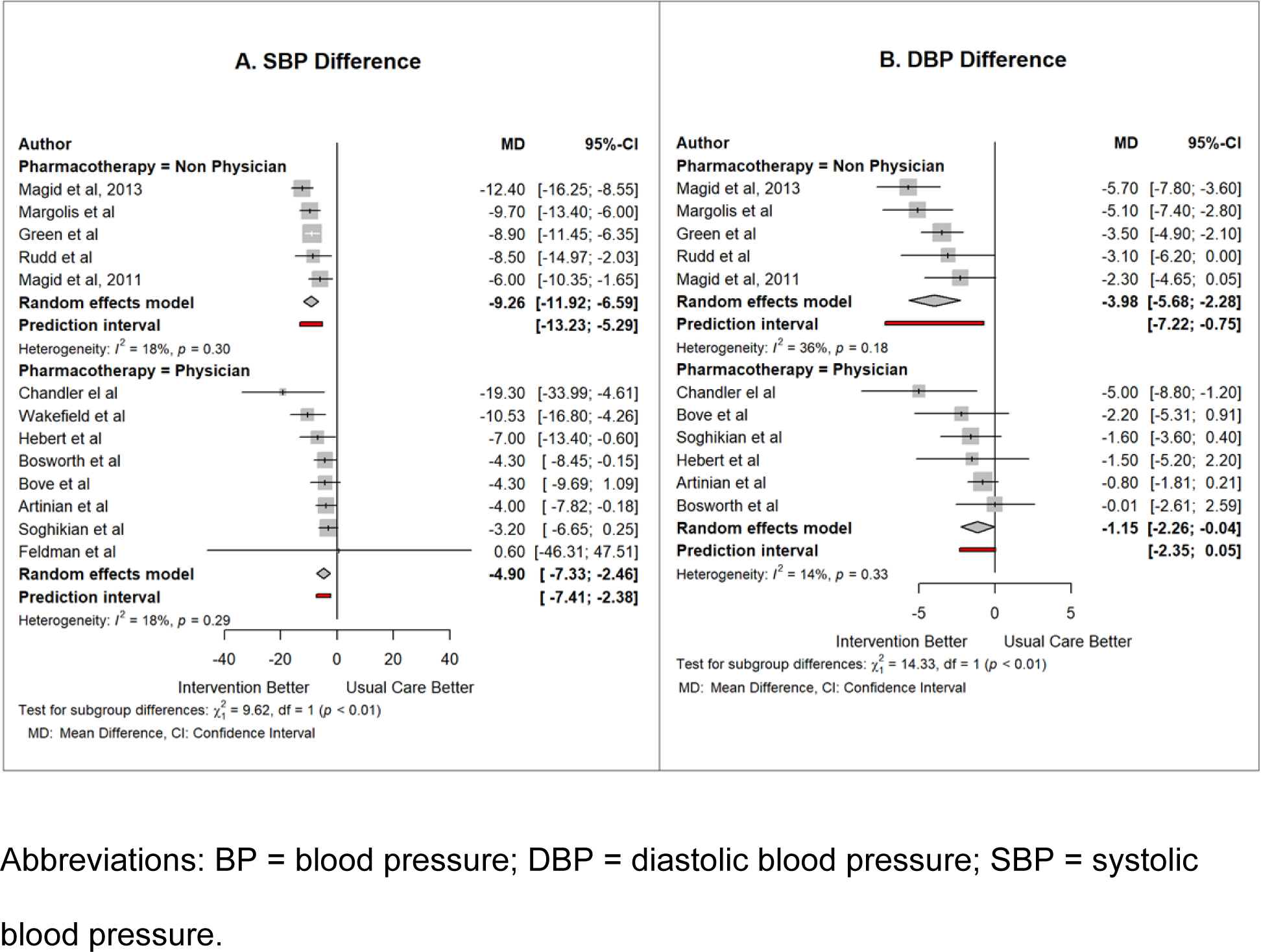
Mean BP difference according to clinicians leading pharmacotherapy.

**Figure 4:**
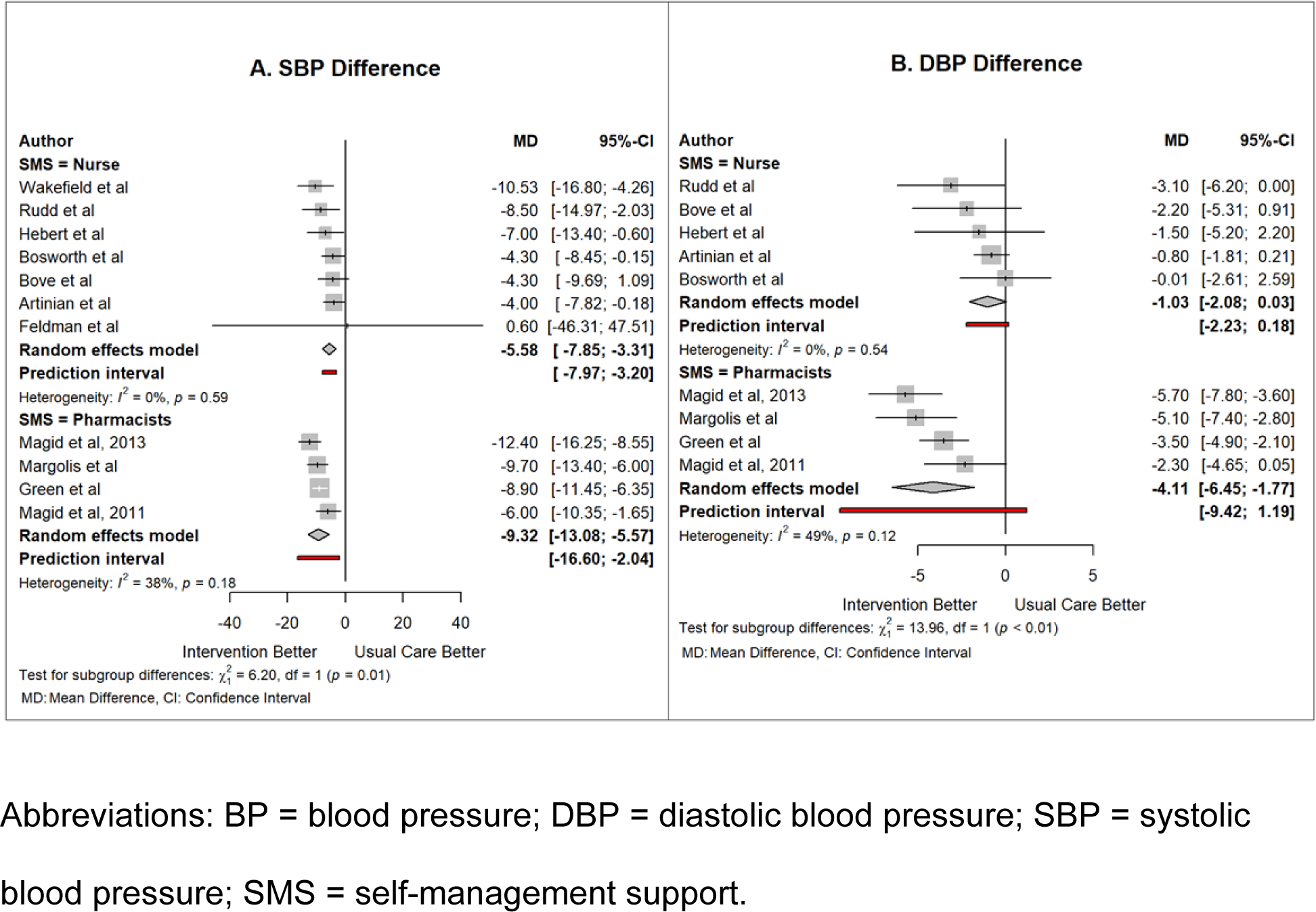
Mean BP difference according to clinicians providing SMS.

**Figure 5:**
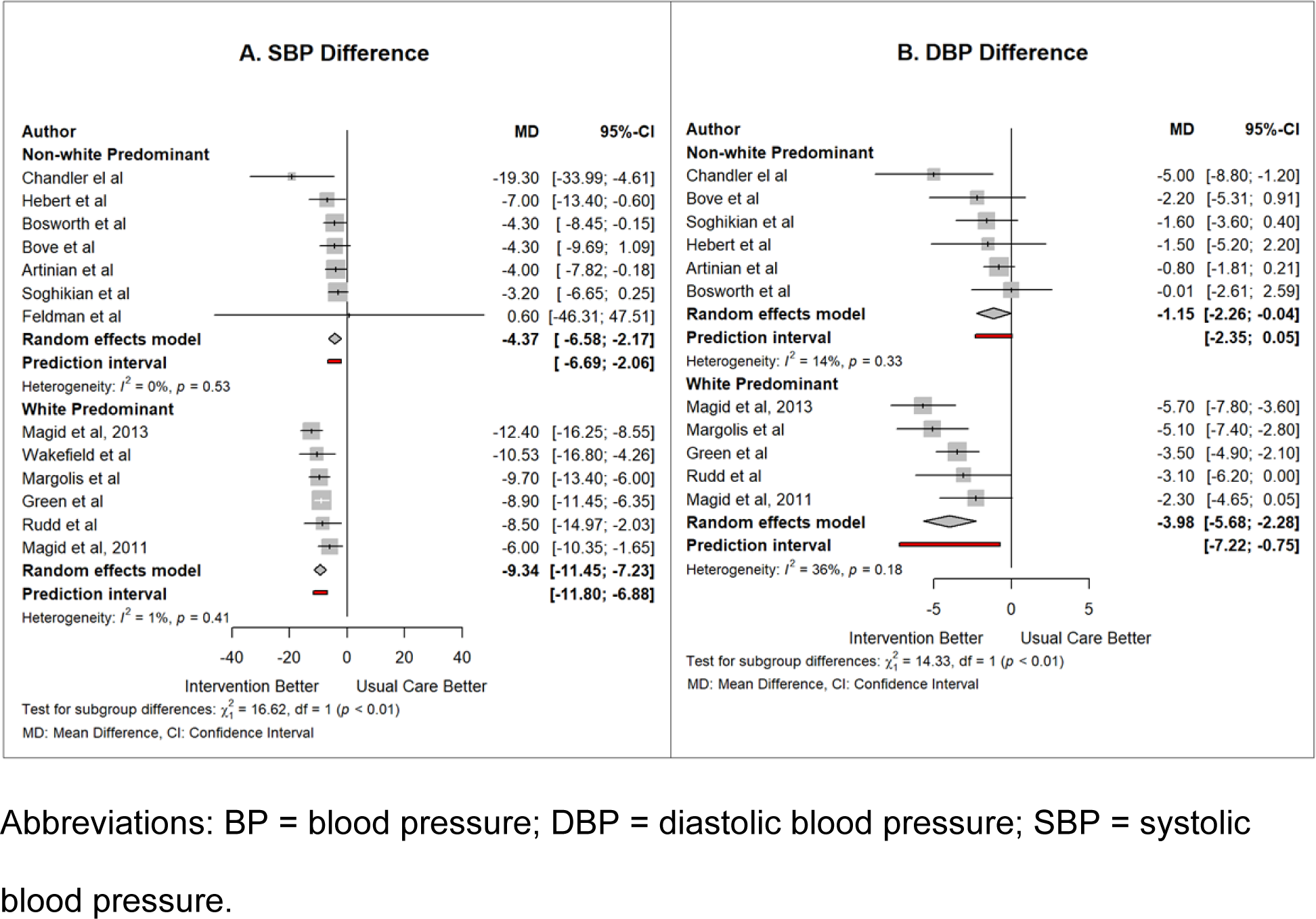
Mean BP difference between majority Non-White vs. White patient predominant trials.

The pooled DBP difference was -2.7 mmHg (95% CI: -4.0, -1.5), favoring TM-HTN intervention (Figure 2). In univariable meta-regression, the only variables significantly associated with DBP difference were clinicians leading pharmacotherapy and SMS, and race. DBP reduction was 2.8 mmHg more when pharmacotherapy was led by non-physicians vs. physicians (P<0.01). Similarly, DBP reduction was 3.1 mmHg more when SMS was provided by pharmacists vs. nurses (P=0.01). For every 10% more non-White patients per trial, DBP reduction was 0.3 mmHg lower (P=0.04) or 2.7 mmHg lower DBP reduction in non-White vs. White patient predominant trials (P<0.01). In subgroup analyses, DBP reductions were -4.0 mmHg (95% CI: -5.7, -2.3) vs. -1.1 (95% CI: -2.3, -0.04) when pharmacotherapy was led by non-physician vs. physician (P<0.01), and -1.0 mmHg (95% CI: -2.1, 0.03) vs. -4.1 mmHg (95% CI: -6.4, -1.8) when SMS was provided by nurses vs. pharmacists (P<0.01), respectively, all favoring TM-HTN (Figures 3-4). Similarly, DBP reductions were -1.1 mmHg (95% CI: -2.3, -0.04) vs. -4.0 mmHg (95% CI: -5.7, -2.3) in non-White vs. White patient predominant trials (P<0.01), respectively, favoring TM-HTN (Figure 5). There were no significant subgroup differences by diabetes predominant trials (P=0.20). Both diabetes and non-White patient predominant trials achieved -0.9 mmHg (95% CI: -1.8, 0.06) DBP reduction vs. - 4.2 mmHg (95% CI: -6.8, -1.6) in White patient predominant but not diabetes predominant trials (P<0.01), favoring TM-HTN.

The mean pooled difference in BP control rate between TM-HTN intervention and usual care was 10.1% (95% CI: 0.4, 19.9), favoring TM-HTN intervention (Figure 2). We did not consider meta-regression and sub-group analysis for BP control due to only seven eligible trials.

To explore the differential effect of SMS on BP, in post-hoc analysis, we conducted sub-group analysis according to whether SMS was well-developed or not. We did not find any significant difference for both SBP [-7.1 mmHg (95% CI: -10.4, -3.9) vs. -8.5 mmHg (95% CI: -12.5, -4.5), (P=0.49)] and DBP [-2.0 mmHg (95% CI: -4.3, 0.4) vs. -3.9 mmHg (95% CI: -5.9, -1.8), (P=0.10)] in trials with and without well-developed SMS, respectively.

To further understand the differences seen by race, in post-hoc analysis, we examined interaction of race with clinicians leading pharmacotherapy, SMS and whether the SMS was well developed or not. We found that the only group achieving lower BP reduction was non-White patient predominant trials (Supplement Table 6). Any group with White patient predominant trials achieved higher BP reduction.

When limiting the analyses to the nine trials that used telemonitoring of SMBP, the pooled SBP and DBP differences were -7.8 mmHg (95% CI: -10.6, -5.0) and -3.0 mmHg (95% CI: -4.7, -1.2), respectively, favoring TM-HTN intervention (Figure 6), which were not significantly different than the four trials without telemonitoring, SBP [-5.3 mmHg (95% CI: -9.6, -0.9), P=0.16] and DBP [-1.9 mmHg (95% CI: -3.9, 0.04), P=0.24], respectively. TM-HTN trials using telemonitoring of SMBP were 13.9% (95% CI: 2.3, 25.5) more likely to control BP than usual care (Figure 6). In univariable meta-regression, the only variable significantly associated with SBP difference was race and for DBP difference was SMS (data not shown). Results from sub-group analyses were similar to the main analyses (data not shown).

**Figure 6:**
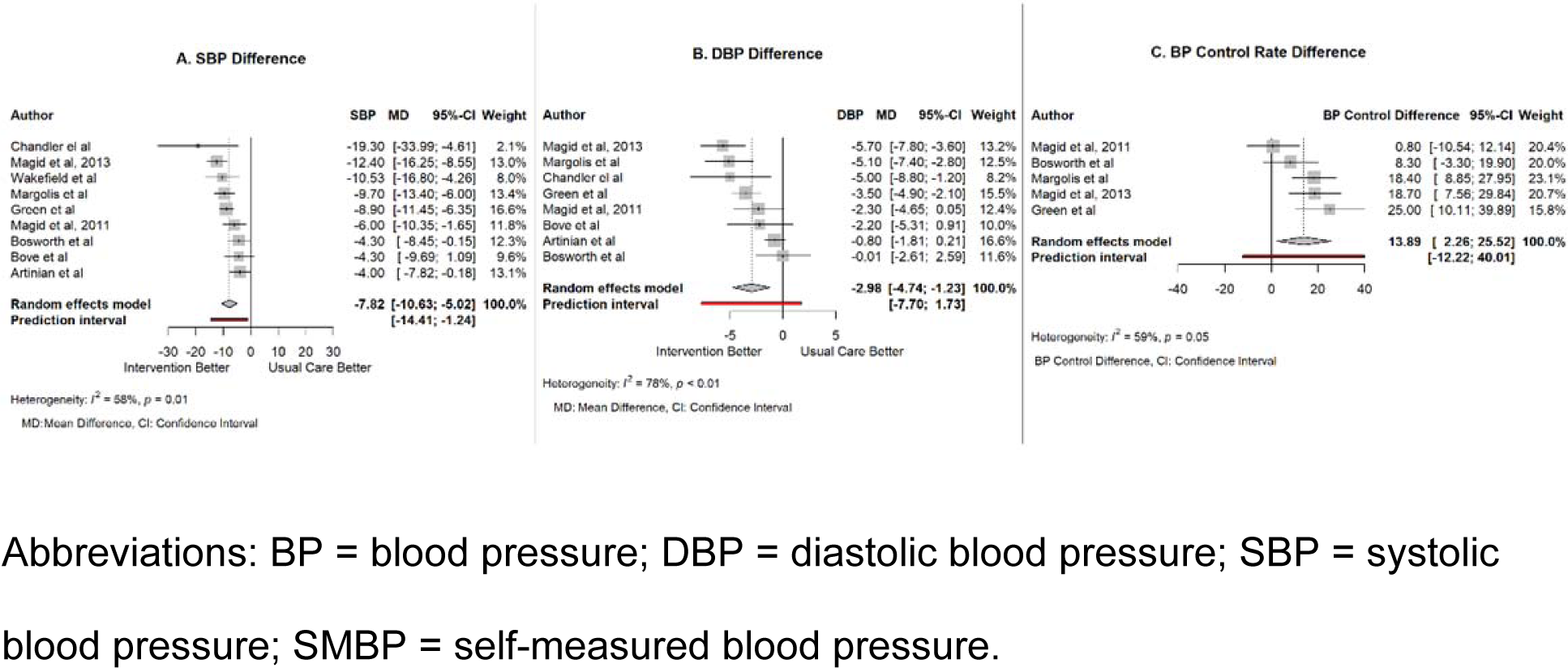
Overall BP outcomes in studies using tele monitoring of SMBP.

## Discussion

To inform translation of TM-HTN in clinical practice in the US, we conducted a systematic review and meta-analysis of US-based randomized trials. We found high strength of evidence for TM-HTN trials lowering BP by 7.3/2.7 mmHg compared with usual clinic-based care, based on direct evidence, with mostly low study limitations and without reporting bias (Supplement Table 5). Pharmacotherapy was more effective if delivered by non-physicians (9.3/4.0 mmHg) than by physicians (4.9/1.1 mmHg), and SMS was more effective when delivered by pharmacists (9.3/4.1 mmHg) than by nurses (5.6/1.0 mmHg). Similarly, TM-HTN was more effective in reducing BP in trials primarily enrolling White patients (9.3/4.0 mmHg) than non-White patients (4.4/1.1 mmHg). There were no subgroup differences by diabetes patient predominant trials. Both diabetes and non-White patient predominant trials achieved lower BP reduction (4.5/0.9 mmHg) than in White patient predominant but not diabetes predominant trials (9.5/4.2 mmHg). Results were similar when limiting the analysis to trials using telemonitoring of SMBP. Taken together, our study provides important insights for effective and equity-conscious TM-HTN implementation in the US (Figure 7).

**Figure 7.**
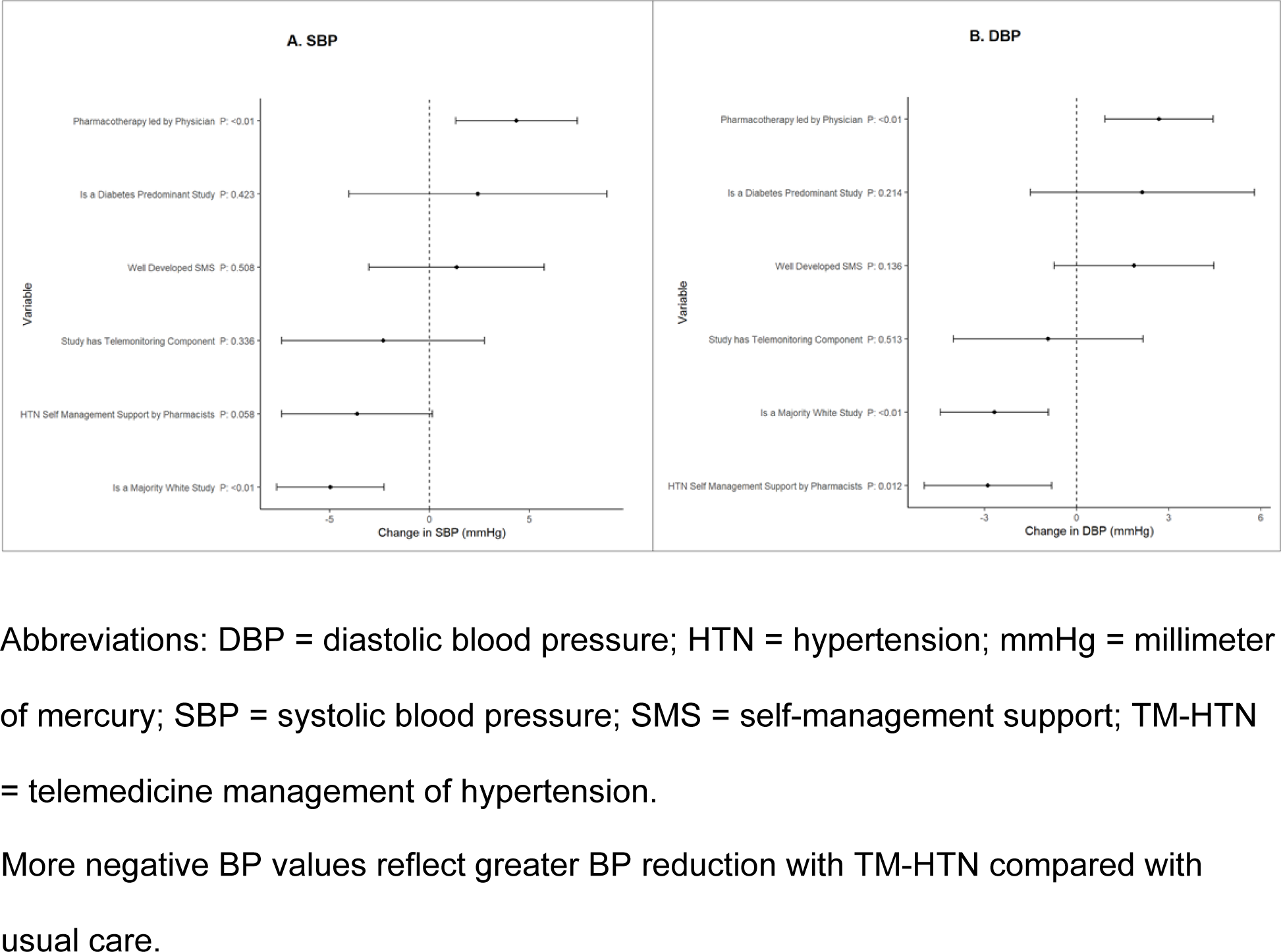
Univariable regression forest plot.

Telemedicine is particularly appropriate for HTN management.^2^ Obtaining accurate and reliable BP is a basic requirement in HTN management. Yet accurate clinic BP evaluation is challenging with limited assessment during sporadic and brief clinic visits,^4^ which undermines clinicians confidence to escalate anti-hypertensive medications.^5,6^ TM-HTN program integrates multiple home BP data, which enhances clinicians’ confidence to assess and manage HTN, and helps overcome clinical inertia.^2^ Enhancing patients’ self-management skills is an important aspect of HTN care.^7^ Self-management comprises patients’ knowledge about HTN, skills in medication adherence, lifestyle modifications, and chronic disease management.^7^ Effective SMS is time consuming requiring ongoing and organized support.^7^ Brief and infrequent clinic visit does not allow effective SMS. TM-HTN is an ongoing, team-based, HTN focused management approach allowing effective SMS.^2^ Taken together, TM-HTN addresses several shortcomings of the clinic-based HTN care.

Multilevel, multicomponent strategies, such as team-based care with medication titration by non-physicians in clinic-based HTN management models are the most effective approaches for BP control.^11-13^ We found similar theme in the telemedicine approach where non-physicians led pharmacotherapy was more effective than when directly led by physicians. It is known that allowing non-physicians to offer pharmacotherapy without needing routine approval from physicians will streamline workflow, offer patient-centered collaborative care, and a focused algorithmic care by non-physicians encourages guideline concordant care while allowing physicians to focus on complex cases. ^37^ In practice, non-physicians providing pharmacotherapy in a telemedicine program for a single condition is a matter of scalability, economy and efficiency. We found similar theme for SMS by pharmacists vs. nurses. Future studies should explore reasons for such difference. Adopting effective TM-HTN programs requires considering these practical considerations and conducting local formative assessment.

Our race pertinent analysis should be cautiously interpreted as the results could be explained by other confounders. For example, we found more diabetes predominant trials and well-developed SMS programs in non-White than White patient predominant trials. Generally, a well-developed SMS intervention is focused on enhancing patient’s self-care skills. Conversely, an intervention without well-developed SMS program may have other focus (e.g., pharmacotherapy). Despite not noting significant BP differences based on well-developed SMS intervention, confounding is still possible. We found no single non-White patient predominant trial that had non-physician leading pharmacotherapy or pharmacists delivering SMS, which were both associated with the highest improvement in BP. Nevertheless, racial differences were notable throughout our analyses. It’s possible that TM-HTN program implemented as such can exacerbate inequities in HTN care in non-White patients. The attenuated BP difference in non-White patient predominant trials (most of whom were Black patients) is likely because the current TM-HTN programs were not designed and tailored to meet these patients’ needs.^23^ For example, Black patients have reported several factors contributing to poor BP control, including limited HTN knowledge, cultural factors, racism-related stress, limited access to community resources and social determinants of health.^38-40^ Structural factors contributing to racial disparities in telemedicine use for diabetes care has been described previously.^41^ Accordingly, non-White patients will need proper support for effective TM-HTN use.^42^ A barber shop-based, pharmacist managed HTN care intervention was highly effective, supporting the importance of a contextually appropriate intervention.^43^ To improve equity in HTN care, TM-HTN programs for non-White patients can adapt the intervention considering patient, clinician, and system-level barriers.^42^ We found that both non-White patient and diabetes predominant trials achieved smaller BP reduction than White patient predominant but not diabetes predominant trials. The additional complexity to address diabetes in non-White population perhaps add to the support needed for optimal use of SMBP. We did not find variation in TM-HTN’s effectiveness by patient’s age, sex, education, diabetes status, baseline BP, and TM-HTN intervention duration.

To our knowledge, this is the first meta-analysis limited to trials using SMBP-guided pharmacotherapy. There are several meta-analyses of trials conducted both in and outside the US using SMBP with or without additional support.^8-10^ The additional support was heterogeneous comprising either SMS only or SMBP-guided pharmacotherapy only or both. SMBP alone can lead to some reductions in SBP (3-5 mmHg) and DBP (2-3 mmHg) at 6 months without difference at 12 months.^8-10,44,45^ When SMBP is accompanied by co-interventions, like SMS, there can be moderate reductions in BP and improved BP control at 12 months.^9^ The SBP difference of 7.3 mmHg seen in our study is comparable to 6.1 mmHg reduction seen in a meta-analysis including trials from outside the US when SMBP was combined with intensive support.^9^ Similarly, a prior meta-analysis found little heterogeneity of the benefit of SMBP across most HTN-related co-morbidities.^22^

Although our study provides important insights into TM-HTN implementation, there are other potential patient, clinician, and system level challenges in optimal use of TM-HTN. For example, uptake of telemedicine intervention was 98% in an efficacy trial of TM-HTN,^28^ but in subsequent pragmatic trial with similar intervention, telemedicine uptake was only 27%,^46^ highlighting the practical challenges in TM-HTN implementation.^47^ Some patient related barriers in using TM-HTN intervention include digital literacy, internet and smart phone access and sociocultural factors.^42,44^ A 2021 survey of nationally representative Americans 50-80 years of age showed that ∼77% of adults with HTN have home BP device with an arm cuff but only ∼25% share their home BP readings with clinicians.^48^ For effective use, patients should receive proper training on SMBP and this training process should be integrated into the overall system of care.^2^ Another survey of primary care clinics showed that only 27.6% of clinics had written policy to train patients in using SBMP and 48.8% designated a team member to provide training to patients.^49^ A pragmatic trial showed that only 38% of patients were able to bring home BP diaries to clinicians, highlighting real-world challenges in using SMBP.^50^ Therefore, understanding and addressing barriers to SMBP and seamlessly transferring home BP values to clinics are important for the success of TM-HTN implementation.^44,50^ In this context, the results seen in our analysis limiting to trials that used telemonitoring of SMBP, which was similar to the main results, should encourage adoption of telemonitoring of SMBP in contemporary TM-HTN programs. Telemonitoring facilitates efficient SMBP communication with clinicians and can expedite clinical decisions.^10^ Clinicians and system related relevant factors include availability of trained personnel to provide SMS, acceptability and readiness among patients and clinicians in adopting a new model of care and having a technical infrastructure to integrate home BP data into electronic medical records. These factors should be explored in future studies.

We caution interpretation of our results considering the following potential limitations. Despite our effort to reduce and address heterogeneity between trials during trials selection and data analysis, some heterogeneity exists. Nevertheless, the confidence interval of each individual trial’s effect size overlapped the point estimate or the confidence interval of the pooled effect, suggesting the plausibility of the variation in effect size among trials.^21^ Our results, particularly the subgroup findings, imply associations and not causations as limited number of eligible trials restricted multivariable meta-regression modeling to address trial-level heterogeneity. One trial had high risk of bias from missing data. Three additional trials had “some concern” for RoB from lack of pre-specified protocols as they were published when reporting protocols were not routine. Although there was heterogeneity in the definition of BP control across trials, the definition mostly reflected contemporary practice guidelines suggesting appropriate practice standards in both intervention and control arms. There were also minor differences in usual care across trials. Finally, non-White patient predominant trials comprised of multiple races with Black race being majority. The attenuated BP effect noted could be related to the differences in the sociocultural factors relevant for multiple races and ethnicities, which is important to consider when implementing TM-HTN intervention.

## Conclusions

TM-HTN is more effective than clinic-based care for BP management in the US. The benefit appears to be pronounced if pharmacotherapy is led by non-physician than physicians and SMS is provided by clinical pharmacists than nurses, although the reason for the latter should be first clarified in future studies. Trials enrolling majority non-White patients seemed to achieve lower BP reduction than trials enrolling majority White patients. As health systems plan to use TM-HTN programs in the US, locally informed, patient-centered adaptation of TM-HTN is needed to provide equitable HTN care.

## Data Availability

All the relevant data will be available on request.

https://doi.org/10.17605/OSF.IO/NHMB4

## Abbreviations

BP: blood pressure
DBP: diastolic blood pressure
HTN: hypertension
RoB: risk of bias
SBP: systolic blood pressure
SMBP: self-measured blood pressure
SMS: self-management support
TM-HTN: telemedicine management of hypertension

